# Association of newly diagnosed atrial fibrillation with remote intracerebral hemorrhage after intravenous thrombolysis: Results from a multicenter study in China

**DOI:** 10.1101/2023.02.22.23286328

**Authors:** Xiaoling Pan, Yingjian Pei, Meixia Zhang, Wansi Zhong, Jin Hu, Zhimin Wang, Dongjuan Xu, Min Lou, HongFang Chen, Zhicai Chen

## Abstract

**Objective:** To investigate the association of atrial fibrillation (AF), especially newly diagnosed AF, with remote intracerebral hemorrhage (rICH) in patients with ischemic stroke treated by intravenous thrombolysis.

**Methods:** This is an observational study of patients with ischemic stroke who were treated with intravenous thrombolysis with recombinant tissue-type plasminogen activator whose data were taken from a multicenter prospective registry of a Chinese population. RICH was defined as any extraischemic hemorrhage detected by imaging examination ≤ 24 hours after intravenous thrombolysis. We collected the demographic data and clinical characteristics of all the patients. We compared patients with rICH and those without any type of hemorrhagic transformation. The association of AF and rICH was analyzed by univariate analysis and binary logistic regression.

**Results:** We evaluated 20697 patients, 1566 (7.6%) of whom developed intracerebral hemorrhage (ICH), 586 (2.8%) of whom experienced rICH, and 19131 (92.4%) of whom did not experience any type of hemorrhagic transformation. Univariate analysis showed that there were significant differences in age, prethrombolysis systolic blood pressure, baseline NIHSS score, known AF, newly diagnosed AF, coronary heart disease, congestive heart failure, hyperhomocysteinemia and history of thrombolysis between the rICH and control groups (P < 0.05). Further multivariate logistic regression analysis showed that total AF [OR 1.821, (95%CI, 1.082-3.065), P < 0.05], known AF [OR 1.470, (95%CI, 1.170-1.847)] and newly diagnosed AF [OR 1.920, (95%CI, 1.304-2.825)] were all independently associated with rICH.

**Conclusions:** This study suggests that AF (regardless of the newly diagnosed or known AF) may be associated with the occurrence of rICH after intravenous thrombolysis. Interestingly, newly diagnosed AF may have a greater impact on rICH than known AF, but that finding needs to be confirmed by a larger prospective sample.

## 1. Introduction

Ischemic stroke is the most common type of cerebrovascular disease with high morbidity, disability, and mortality. There is a global consensus that intravenous injection of recombinant tissue plasminogen activator is one of the most effective methods for the treatment of ischemic stroke within the time window and that it can greatly improve the outcomes of patients. However, intracerebral hemorrhage (ICH) is the most severe complication of intravenous thrombolysis (IVT) for acute ischemic stroke. The majority of ICH occurs within or at the margin of infarcted brain tissue, and is classified as an intracranial hemorrhage transformation (HT)^1^. Sometimes, ICH may appear in regions without visible ischemic damage, which is defined as remote intracerebral hemorrhage (rICH) in the Heidelberg classification^2^. In addition to HT, the presence of rICH is also associated with worse functional outcomes^3-5^ and a higher risk of mortality^6^. Therefore, the investigation of the risk factors related to rICH in ischemic stroke patients after IVT may be clinically important because it may indicate potential prevention methods.

Atrial fibrillation (AF) is one of the most common risk factors for ischemic stroke, accounting for approximately 15-20% of cases^7^. According to recent guidelines, AF-associated strokes can benefit from IVT^8^. Previous studies have demonstrated that AF is associated with HT after IVT^9^. There is less literature on the relationship between AF and rICH, and the relationship between AF and rICH has not been fully clarified^3, 10^. Previous studies showed that among acute ischemic stroke patients with AF, 7.8 to 36.2% were newly diagnosed with AF after stroke^11-13^. Patients with newly diagnosed AF after stroke and those with known AF before stroke have different background characteristics. To the best of our knowledge, no previous study has explored the relationship between different types of AF (known AF and newly diagnosed AF) and rICH. Therefore, the purpose of this study was to investigate the relationship between different types of AF and rICH after IVT. This study retrospectively analyzed the clinical data of a multicenter, large sample of patients with IVT in Zhejiang Province, China, to investigate the association of AF with rICH in patients with ischemic stroke after IVT.

## 2. Methods

### 2.1 Study population

This is a multicenter, prospective, observational study of patients with ischemic stroke treated with IV recombinant tissue plasminogen activator(rt-PA) at 71 different hospitals in Zhejiang Province, China, between June 2017 and December 2021. All patients were prospectively and consecutively included in the Zhejiang Stroke Medical Quality Control Center Platform, which is an academic institution based at the Second Affiliated Hospital of Zhejiang University School of Medicine to monitor the quality of all reperfusion therapy cases performed at the stroke centers in Zhejiang Province, China. We included acute ischemic stroke patients aged ≥ 18 years who were treated with IV rt-PA within the first 4.5 hours from the onset of symptoms. Exclusion criteria included patients who had no documented brain CT/MR results within the first 24 hours after thrombolytic therapy and patients who underwent any modality of endovascular therapy. The following variables were recorded: (1) demographics (age, sex); (2) traditional vascular risk factors [smoking, hypertension, diabetes mellitus, hyperlipidemia, previous ischemic stroke, AF, coronary heart disease (CHD), congestive heart failure (CHF) and hyperhomocysteinemia (Hcy)]; (3) previous medications (antiplatelet agents, anticoagulants, and statins); (4) blood pressure before thrombolysis; (5) onset to treatment time (OTT); (6) severity of the neurological deficit as measured by the National Institutes of Health Stroke Scale (NIHSS) score; and (7) etiology of stroke [TOAST (Trial of Org 10172 in Acute Stroke Treatment) classification]. Intravenous thrombolytic therapy with rt-PA (0.9 mg/kg up to a maximum of 90 mg) was used with 10% of the total dosage as a bolus, and the rest was infused over 1 hour.

This study was reviewed by the Ethics Committee of the Second Affiliated Hospital of Zhejiang University School of Medicine (2016 Annual Ethics Review No. 074), and all patients and their family members gave informed consent for relevant examination and treatment.

### 2.2 Definition of AF

All the patient ‘s past medical history was recorded, along with an electrocardiogram and continuous ECG monitoring for 24 to 48 hours. AF was classified as known AF and newly diagnosed AF. Known AF was defined when the previous history of AF was provided by patients or medical recording systems. Newly diagnosed AF was defined when there was no previous history of AF, but AF was detected on ECG or ECG monitoring during the current visit, regardless of the emergency department or neurology department during this hospital stay.

### 2.3 Definition of HT and rICH

In this study, HT was defined as a new hemorrhage within infarcted brain tissue. HT was classified according to the European Cooperative Acute Stroke Study criteria^1^, including hemorrhagic infarct (HI) and parenchymal hemorrhage (PH). HI was further divided into HI1 (small petechiae) and HI2 (confluent petechiae), and PH was divided into PH1 (<30% of the infarcted area with mild space-occupying effect) and PH2 (> 30% of the infarcted area with obvious mass effect). RICH was defined as ICH in a brain region without visible ischemic damage on 24-hour CT/MR scans according to the Heidelberg bleeding classification^2^.

### 2.4 Statistical analysis

We divided the patients according to bleeding complications. RICH consisted of rICH patients alone and rICH combined with HI/PH. Patients without ICH comprised the no-bleeding group, which was the control group.

We compared demographic variables, traditional vascular risk factors, previous medications, baseline NIHSS, blood pressure before thrombolysis, OTT, and stroke pathogenesis between patients from the rICH group and patients from the control group. The normally distributed measurement data were expressed as the means ± SD, and the average values between the two groups were compared using the t test. Data with nonnormal distributions were expressed as medians (25th-75th percentile) [M (Q1, Q3)], and pairwise comparisons were performed using the Mann–Whitney U test. Categorical data were expressed as numbers and percentages and were compared using the χ^2^ test. The level of statistical significance was defined as a two-sided test. Variables with P < 0.05 in univariate analysis were entered into the multivariate logistic regression analysis model. All statistical analyses were performed using SPSS, Version 22.0 (IBM, Armonk, New York). P < 0.05 was considered statistically significant.

## 3. Results

A total of 24405 patients with ischemic stroke were treated with reperfusion therapy (including IVT and endovascular therapy) within the study period. We excluded 1881 patients without a follow-up noncontrast CT within the first 24 hours of IV-rt-PA and 1827 patients who underwent endovascular therapy. Finally, 20697 patients were enrolled (Figure 1). Among 20697 patients, 1566 (7.6%) patients had ICH, and 19131 (92.4%) patients did not have any type of intracranial hemorrhage transformation (Table 1).

**Table 1.** Distribution of Bleeding Complications

|  |  |
| --- | --- |
| No-ICH | 19131 (92.4%) |
| HT without rICH | 980 (4.7%) |
| HI1 | 234 (1.1%) |
| HI2 | 388 (1.9%) |
| PH1 | 187 (0.9%) |
| PH2 | 171 (0.8%) |
| rICH | 586 (2.8%) |
| isolate rICH | 358 (1.7%) |
| rICH concomitant HT | 228 (1.1%) |
| Total | 20697 |

**Figure 1.**
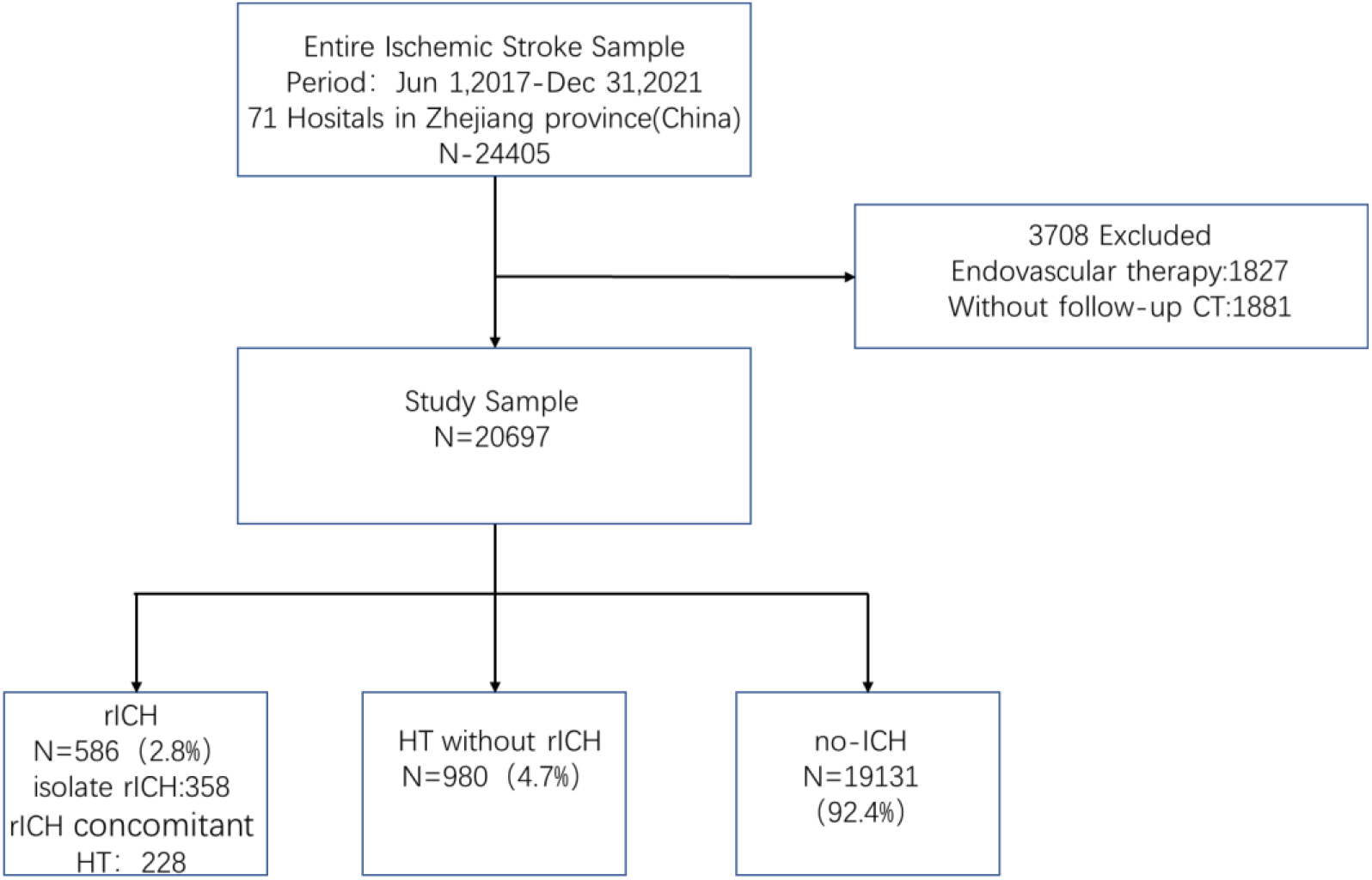
Study flow diagram.

### 3.1 Results of univariate analysis between the rICH group and the control group

Table 2 shows the results of a univariate analysis of baseline demographics, vascular risk factors, previous medications, and related indicators before thrombolytic therapy between the rICH group and the no-ICH group. Compared with the control group, patients in the rICH group were older (74 vs. 69 years, P < 0.001) and had higher systolic blood pressure before thrombolytic therapy (156 vs. 153 mmHg, P = 0.002), higher baseline NIHSS (7 vs. 4, P < 0.001), higher rate of total AF (29.9% vs. 15.9%, P < 0.001), higher known AF (24.6% vs. 13.5%, P < 0.001) and newly diagnosed AF (5.3% vs. 2.4%, P < 0.001), higher history of CHD (9.9% vs. 7.6%, P = 0.038), higher history of CHF (4.4% vs. 1.9%, P < 0.001), higher Hcy (9.9% vs. 6.6%, P = 0.002), higher history of thrombolysis (2.7% vs. 1.4%, P = 0.008) and a higher rate of cardiac cerebral embolism (31.9% vs. 16.4%, P < 0.001).

**Table 2.** Bivariate analyses of baseline variables for patients with rICH and patients without any type of ICH

| Variables | rICH (n = 586) | No-ICH (n = 19131) | t/X<sup>2</sup> | P |
| --- | --- | --- | --- | --- |
| Age, y, mean ± SD | 74 ± 11.95 | 69 ± 12.76 | 9.398 | 0.000 |
| Male, n (%) | 356 (60.8) | 12075 (63.1) | 1.367 | 0.242 |
| **Risk factors and previous diseases** |  |  |  |  |
| Smoking, n (%) | 157 (26.8) | 5814 (30.4) | 3.488 | 0.062 |
| Hypertension, n (%) | 394 (67.2) | 12337 (64.5) | 2.702 | 0.100 |
| Diabetes mellitus, n (%) | 83 (14.2) | 3201 (16.7) | 1.032 | 0.310 |
| Hyperlipidemia, n (%) | 27 (4.6) | 1138 (5.9) | 1.839 | 0.175 |
| Hcy, n (%) | 58 (9.9) | 1258 (6.6) | 10.073 | 0.002 |
| Previous ischemic stroke, n (%) | 82 (14.0) | 2431 (12.7) | 0.846 | 0.358 |
| History of thrombolysis, n (%) | 16 (2.7) | 268 (1.4) | 7.079 | 0.008 |
| AF, n (%) | 175 (29.9) | 3034 (15.9) | 81.835 | 0.000 |
| Known AF, n (%) | 144 (24.6) | 2575 (13.5) | 59.071 | 0.000 |
| Newly diagnosed AF, n (%) | 31 (5.3) | 459 (2.4) | 20.293 | 0.000 |
| CHD, n (%) | 58 (9.9) | 1451 (7.6) | 4.304 | 0.038 |
| CHF, n (%) | 26 (4.4) | 366 (1.9) | 18.585 | 0.000 |
| **Previous treatments** |  |  |  |  |
| Antiplatelets, n (%) | 89 (15.2) | 2606 (13.6) | 0.277 | 1.181 |
| Anticoagulants, n (%) | 10 (1.7) | 261 (1.4) | 0.491 | 0.483 |
| Statins, n (%) | 67 (11.4) | 1828 (9.6) | 2.309 | 0.129 |
| Systolic blood pressure, mmHg, mean ± SD | 156 ± 22.00 | 153 ± 21.16 | 3.123 | 0.002 |
| Diastolic blood pressure, mmHg, mean ± SD | 85 ± 13.65 | 85 ± 13.44 | 0.895 | 0.371 |
| OTT, min, median (IQR) | 154 (109,202) | 146 (105,193) | -1.856 | 0.064 |
| NIHSS baseline, median (IQR) | 7 (3,14) | 4 (2,9) | -9.410 | 0.000 |
| **TOAST** |  |  |  |  |
| Cardiocerebral embolism, n (%) | 187 (31.9) | 3144 (16.4) |  | 0.000 |
| Noncardiac cerebral embolism, n (%) | 399 (68.1) | 15987 (83.6) | 97.009 |  |

### 3.2 Multivariate analysis results of rICH after IVT in patients with ischemic stroke

As shown in Figure 2, after adjustment for covariance using multivariate logistic regression analysis, known AF [OR 1.470, (95%CI, 1.170-1.847)], newly diagnosed AF [OR 1.920, (95%CI, 1.304-2.825)], history of thrombolysis [OR 1.821, (95%CI, 1.082-3.065)], age [OR 1.021, (95%CI, 1.013-1.029)], history of congestive heart failure [OR 1.716, (95%CI, 1.101-2.676)], systolic blood pressure before thrombolysis [OR 1.005, (95%CI, 1.001-1.009)] and baseline NIHSS score [OR 1.027 (95%CI, 1.016-1.038)] were independently associated with the occurrence of rICH.

**Figure 2.**
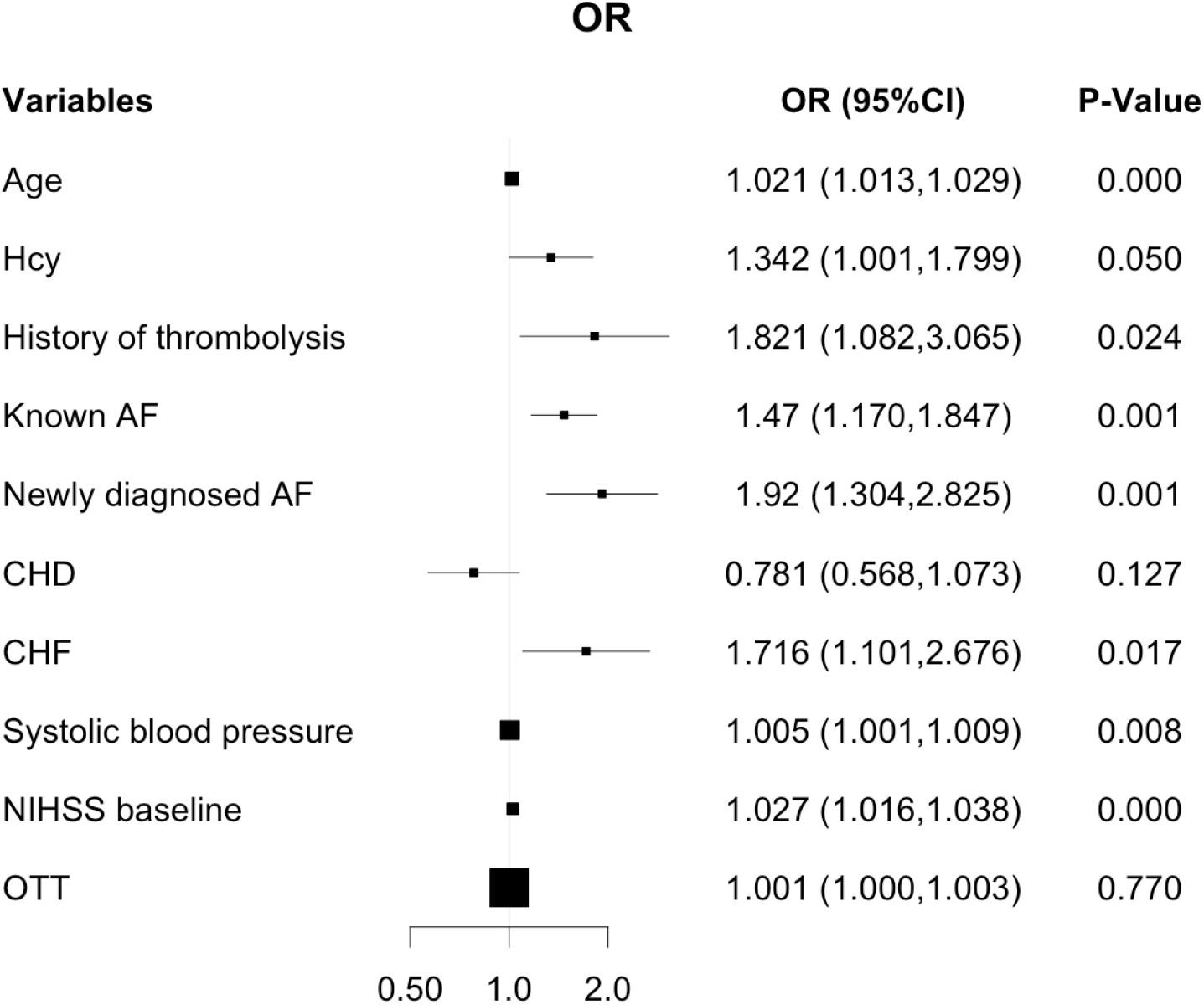
Logistic regression analysis of the risk factors for rICH.

### 3.3 Results of univariate analysis between the newly diagnosed AF group and the known AF group

Table 3 shows the results of univariate analysis between the newly diagnosed AF group and the known AF group. Compared with the known AF group, the newly diagnosed AF group had a higher rate of males (58.2% vs. 52.0%, P = 0.012), a higher rate of smokers (25.3% vs. 20.4%, P = 0.014), and a higher rate of Hcy (11.4% vs. 8.2%, P = 0.018), and the systolic blood pressure (153 mmHg vs. 151 mmHg, P = 0.043) and diastolic blood pressure (86 mmHg vs. 85 mmHg, P = 0.013) before thrombolysis were higher than those in the known AF group. However, previous ischemic stroke (13.3% vs. 17.4%, P = 0.024), history of thrombolysis (1.2% vs. 3.1%, P = 0.021), CHD (9.4% vs. 23.6%, P < 0.001), and CHF (2.2% vs. 9.5%, P < 0.001) were significantly lower than those in the known AF group. Meanwhile, history of antiplatelet therapy (13.5% vs. 26.7%, P < 0.001) and statin therapy (11.0% vs. 16.6%, P = 0.002) were also significantly lower than those in the known AF group.

**Table 3.** Bivariate analyses of the newly diagnosed AF group and known AF group

| Variables | Newly Diagnosed AF (n = 490) | Known AF (n = 2719) | t/X<sup>2</sup> | P |
| --- | --- | --- | --- | --- |
| Age, y, mean ± SD | 76 ± 10.74 | 77 ± 9.58 | 1.456 | 0.146 |
| Male, n (%) | 285 (58.2) | 1415 (52.0) | 6.246 | 0.012 |
| **Risk factors and previous disease** |  |  |  |  |
| Smoking, n (%) | 124 (25.3) | 554 (20.4) | 6.058 | 0.014 |
| Hypertension, n (%) | 331 (67.6) | 1835 (67.5) | 0.001 | 0.978 |
| Diabetes mellitus, n (%) | 59 (12.0) | 362 (13.3) | 0.590 | 0.442 |
| Hyperlipidemia, n (%) | 25 (5.1) | 150 (5.5) | 0.138 | 0.710 |
| Hcy, n (%) | 56 (11.4) | 222 (8.2) | 5.589 | 0.018 |
| Previous ischemic stroke, n (%) | 65 (13.3) | 473 (17.4) | 5.077 | 0.024 |
| History of thrombolysis, n (%) | 6 (1.2) | 84 (3.1) | 5.297 | 0.021 |
| CHD, n (%) | 46 (9.4) | 642 (23.6) | 49.871 | 0.000 |
| CHF, n (%) | 11 (2.2) | 257 (9.5) | 28.175 | 0.000 |
| **Previous treatments** |  |  |  |  |
| Antiplatelets, n (%) | 66 (13.5) | 725 (26.7) | 38.918 | 0.000 |
| Anticoagulants, n (%) | 0 | 222 (8.1%) | / | / |
| Statins, n (%) | 54 (11.0) | 450 (16.6) | 9.589 | 0.002 |
| Systolic blood pressure, mmHg, mean ± SD | 153 ± 22.66 | 151 ± 21.00 | -2.025 | 0.043 |
| Diastolic blood pressure, mmHg, mean ± SD | 86 ± 14.56 | 85 ± 13.75 | -2.473 | 0.013 |
| OTT, min, median (IQR) | 154 (109,202) | 146 (105,193) | -0.557 | 0.578 |
| NIHSS baseline, median (IQR) | 9 (4,16) | 9 (4,16) | 1.405 | 0.160 |

## 4. Discussion

The incidence of rICH is lower than that of HT. Approximately 2.8% of patients developed rICH in our study, which is consistent with previous studies. The National Institute of Neurological Disorders and Stroke of the United States found that the incidence of rICH was 1.3%^14^. The European Collaborative Acute Stroke Study (ECASS) and ECASS II study found that the incidence of rICH was 3.7% and 2.0%, respectively^15^. This is similar to previous findings, suggesting that rICH is uncommon but not rare.

Previous studies have shown that AF is associated with the occurrence of HT^9^, which may be related to factors such as AF easily causing acute occlusion of large arteries, poor collateral circulation, and prolonged recanalization time^16, 17^. AF was associated with PH but not rICH in previous studies, while the study by Prats-Sánchez L et al.^10^ showed that AF was associated with rICH. In our study, AF was an independent risk factor for rICH. The mechanism by which AF increases the risk of rICH is not yet clear. Based on a review of the available literature, we speculate that the possible factors are as follows. First, patients with AF tend to have a history of previous stroke or recent silent brain infarction, while these preexisting vascular lesions have been shown to be associated with rICH^4^. A small prospective study by Drelon A et al.^4^suggested that 41% (17/41) of rICH occurred in previous lesions confirmed by MR before thrombolysis. Second, in patients with AF, previous antiplatelet and anticoagulant therapy may increase the deposition of amyloid in the brain^18-21^, which in turn increases the occurrence of rICH after thrombolysis^22, 23^. Third, AF-associated cardioembolism is associated with an increased burden of small vessel disease burdens [e.g., white matter hyperintensities (WMHs) and cerebral microbleeds (CMBs)]^24^, which were significantly associated with a higher risk of rICH in previous studies^4, 23, 25^.

In this study, approximately 15.3% of AF patients (490/3209, Table 3) were newly diagnosed. Newly diagnosed AF (OR 1.920) seems to have a more significant effect on rICH than known AF (OR 1.470). Our data showed that the patients in the newly diagnosed AF group were more likely to be male and smokers and had higher systolic and diastolic blood pressures before thrombolysis than the known AF group. However, their prevalence of cardiac and cerebrovascular diseases and their rate of receiving antiplatelet, anticoagulant, and statin therapy was lower than the known AF group. To date, there is no clear explanation for this association. It is well known that the risk of asymptomatic cerebral infarction in patients with known AF is reduced by antiplatelet, anticoagulant and statin therapy^26^. Similarly, in studies of newly diagnosed AF, a significant decrease in the risk of embolic events was observed once antiplatelet or anticoagulant therapy was administered^27, 28^. Based on these findings, we hypothesized that patients with newly diagnosed AF were more likely to have a recent silent brain infarction before treatment than those with known AF, especially those who had not received any antithrombotic therapy. As we mentioned above, recent silent brain infarction was associated with rICH^4^. Our results suggest that acute ischemic stroke combined with newly diagnosed AF receiving intravenous thrombolytic therapy has a higher risk of rICH; this association should be given more attention in clinical practice. In this study, 284 patients had received at least two thrombolyses, and a history of thrombolysis [OR 1.821, (95%CI, 1.082 to 3.065)] may be an independent risk factor for rICH. Most of the previous literature on repeated thrombolysis consists of case reports or small sample case series, showing that repeated thrombolysis has no significant effect on the occurrence of HT after IVT^29, 30^. There are no published studies on the effect of previous thrombolytic therapy on rICH after repeated thrombolysis.

Previous studies have observed that intravenous thrombolytic therapy itself could increase intracranial microbleeds^31, 32^, which were associated with the development of rICH after intravenous thrombolytic therapy^33^. In addition, preexisting BBB disruption may be exacerbated after intravenous thrombolysis with alteplase^34^, subsequently prompting rICH. The exact mechanism is still unclear and needs further study.

In addition, our results also showed that age, history of CHF, NIHSS before thrombolysis, and systolic blood pressure before thrombolysis were independently associated with rICH, while OTT were not associated with rICH.

There are limitations to our study. First, the study was a retrospective case−control study even though the relevant information collection and registration were prospective. Second, imaging features before and after thrombolysis were not included in this study, such as high-density shadow of the middle cerebral artery on CT, previous damage lesions, CMBs, white matter lesions, etc. These deletions make it difficult for us to explore the mechanism behind the influence of factors such as AF and previous thrombolysis on rICH. Therefore, these additional factors are worthy of further study.

## 5 Conclusions

In this study, we confirmed that AF (both newly diagnosed AF and known AF) was independently associated with rICH, and newly diagnosed AF had a greater impact on rICH than known AF. These findings may be helpful for selecting patients for future studies to improve the safety of IVT.

## Data Availability

The data underlying this article will be shared on reasonable request to the corresponding author.

AF: atrial fibrillation
rICH: remote intracerebral hemorrhage
ICH: intracerebral hemorrhage
IVT: intravenous thrombolysis
HT: hemorrhage transformation
rt-PA: recombinant tissue plasminogen activator
CHD: coronary heart disease
CHF: congestive heart failure
Hcy: hyperhomocysteinemia
OTT: onset to treatment time
NIHSS: National Institutes of Health Stroke Scale
TOAST: Trial of Org 10172 in Acute Stroke Treatment
ECG: electrocardiogram
HI: hemorrhagic infarct
PH: parenchymal hemorrhage
WMHs: white matter hyperintensities
CMBs: cerebral microbleeds;

## 6 Acknowledgments

We thank all participating hospitals, their physicians and nurses.

## 7 Author Contributions

ZC and HC led the conception and design of the trial and reviewed the manuscript. XP wrote the manuscript and involved in the design of the trial. YP was closely involved in the data collection and data curation and was responsible for the statistical analyses. ML, MZ and WZ were involved in the project administration and supervision. JH, ZW, and DX were involved in the design of the study, participated in the data interpretation, and revised the manuscript critically for important intellectual content.

